# Microbiome evaluation revealed salivary dysbiosis in addicts of betel nut preparations

**DOI:** 10.1101/2020.04.13.20064063

**Authors:** Faizan Saleem, Ghulam Mujtaba, Junaid Ahmed Kori, Arshad Hassan, M. Kamran Azim

**Affiliations:** Department of Biosciences, Mohammad Ali Jinnah University, Karachi, Pakistan; H.E.J. Research Institute of Chemistry, International Center for Chemical and Biological Sciences, University of Karachi, Karachi, Pakistan; Dow Dental College, Dow University of Health Sciences, Karachi, Pakistan

**Keywords:** Oral microbiome, betel nut, Next generation sequencing, metagenomics, 16S rRNA

## Abstract

Betel nut addiction is recognized as the causative agent of oral microbiome dysbiosis and other systematic disorders. A number of betel nut preparations containing ingredients such as slaked lime, catechu extract and tobacco are being commonly used particularly in South Asia. The underlying variations in the oral microbiome due to usage of betel nut preparations are poorly understood. We evaluated salivary microbiome in response to chewing of betel nut preparation(s). In order to assess the microbiome dynamics, metagenomic analysis of 16S rRNA gene (V3-V4 hypervariable region) from salivary bacteria in chewers of betel nut preparation (n = 16) and non-chewers (n = 55) was carried out by Greengenes and SILVA ribosomal sequence databases. It was observed that Gutka chewers demonstrated lower alpha diversity and number of bacterial genera than the non-chewers. Taxonomic assignment on phylum level revealed Firmicutes (p-value = 0.042 at 95% confidence interval) to be significantly more abundant in Gutka chewers in comparison with non-chewers. Beta diversity analysis at genus level by weighted unifrac distance matrices unveiled both groups to be divergent from each other. On the genus level, *Veillonella* (p-value = 0.015), Streptococcus (p-value = 0.026), *Leptotrichia* (p-value = 0.022) and *Serratia* (p-value = 0.022) species appeared to be significantly more abundant in Gutka chewers in comparison to non-chewers. The present study suggests salivary dysbiosis in response to gutka chewing and concludes that gutka chewers possess higher abundance of acidogenic and aciduric bacteria. This study contributes additional information regarding oral microbiome variations with response to gutka consumption.

## 1. Introduction

Betel nut consumption is practiced by 10-20% of the world’s total population and commonly associated with Asian-pacific region^1^. Betel nut (also known as areca nut) has been classified as a group I carcinogen by international agency of cancer research (IARC) and is proposed to generate dependency in consumers^2^. Components of Betel nut have previously been correlated with pro-carcinogenic variations, reactive oxygen species production and immuno-modulatory disruptions^3^. Hence, betel nut chewing is detrimental for oral health by inducing inflammatory responses and periodontal disorders^4^.

Asia pacific region is Hub for a variety of betel nut preparations. In addition to betel nut, the preparations are comprised of slaked lime, catechu, spices and tobacco^5^. Among these preparations ‘Gutka’ (containing betel nut, lime, catechu, sandalwood and tobacco), “Mawa” (containing betel nut, tobacco and slaked lime) and “Paan” (betel nut, slaked like, catechu extract wrapped in piper leaf with or without tobacco) are common in Indian subcontinent^6,7^. According to the epidemiological studies carried out in past few decades, ∼20-40% population of Pakistan, Nepal and India consume these preparations^8^.

Habitual users of gutka experience higher periodontal inflammation, gum bleeding and marginal bone loss that lead to a number of diseases including oral submucosal fibrosis, dental caries and oral cancer^9^. Ingredient of Gutka preparations have been reported to be involved in oral health impairment. Slaked lime promotes production of reactive oxygen species and betel nut extract disrupts the functionality of periodontal fibroblasts, thus leading to increased rate of inflammation and carcinogenesis^9^.

The human oral microbiome is composed of ∼700 different bacterial genera among which 32% phylotypes are unculturable^10,11^. These bacterial communities play a role in maintaining normal oral homeostasis including defense against pathogens, inflammatory response regulation and neutralization of reactive nitrogen species^12^. Several studies have proclaimed the prominence of oral microbiome in development of oral systematic disorders such as gingivitis, dental caries and periodontitis^13^. A number of factors such as lifestyle habitats and tobacco usage can alter the oral microbiome^14^. Consumption of betel nut preparations is among the factors that are responsible for dysbiosis of oral microbiome, thus disrupting the normal oral homeostasis^15^.

However, the alteration of oral microbiome in response to consumption of betel nut preparations such as Gutka is poorly understood. The objective of present study was salivary microbiome evaluation in addicts of betel nut preparations.

## 2. Materials and Methods

### 2.1. Study Population and Sample Collection

The study was sanctioned by Independent Ethics Committee, International Center for Chemical and Biological Sciences, University of Karachi, Pakistan (Study no. 011-SS-2016. Protocol no. ICCBS/IEC-011-SS-2016/Protocol/1.0). The studied population included individuals residing in Karachi, Pakistan within the age limit of 25-40. Studied participants were recruited in the interval of two years from January 2017 to December 2018. Verbal screening of participants was carried out and written informed consent was obtained. In brief, participants chewing Gutka for more than one year with a chewing frequency of at least 5 times/day and consumption of at least 20 gram/day were selected. For non-chewers (n = 55), the individuals having un-controlled HbA1c levels (i.e. HbA1c > 6.0), acute or chronic illnesses, periodontal diseases, tobacco usage, pregnancy and antibiotic usage were not included in the study. For the gutka chewers (n = 16), individuals having current infectious conditions (i.e. white cell count > 11), smoking habit, pregnancy, medication usage, cancerous malignancies, immunosuppression or immunodeficiency treatment and usage of any form of steroids were excluded from the study. HbA1c estimation of each participant was carried out by Hitachi 902 auto-analyzer. Unstimulated saliva samples were obtained from participants in sterilized 15 mL conical tubes and stored at -20 °C till the extraction of bacterial DNA.

### 2.2. DNA Extraction and PCR Amplification

Metagenomic DNA was extracted from saliva samples by using ORAgene DNA extraction kit (DNA Genotek Inc, Ontario, Canada). Quality assessment of extracted DNA was carried out by 1% agarose gel electrophoresis. DNA quantification was performed by Qubit fluorimeter 2.0 (Invitrogen Inc. USA). The V3-V4 hypervariable region of 16S rRNA gene was amplified according to the instructions in ‘16S Metagenomics Sequencing Library Preparation guidelines’ (Illumina Inc. USA) by T100 thermal cycler (BioRad, USA).

### 2.3. 16S rDNA Library Preparation and Sequencing

16S rDNA amplicons were used for index PCR by Nextera XT index kit (Illumina Inc., USA). A final reaction of 50 µl was prepared with 5 µl amplicon, 5 µl index primer 1, 5 µl index primer 2, 25 µl 2X KAPA Hot-Start master mix and 10 µl PCR grade water. Index PCR program was set as, preheating for 3 minutes at 95°C, 8 cycles of initial denaturation at 95°C for 30 seconds, annealing for 30 seconds at 55 °C, extension for 30 seconds at 72°C and final extension for 5 minutes at 72°C. Index PCR clean-up was carried by AMPure XP magnetic beads (Beckman Coulter Inc., USA) according to instructions in ‘16S Metagenomics Sequencing Library Preparation’ guidelines. Cleaned indexed amplicons were diluted to 4nmol concentration followed by pooling. Final pooled library was sequenced by Illumina MiSeq system using V3 reagent kit (Illumina Inc., USA).

### 2.4. Bioinformatics and Statistical Analysis

MiSeq sequencing resulted in 2X300 nts paired-end reads. Quality assessment of paired-end NGS reads was carried out by FASTQC software (Andrews, 2010). NGS reads with poor quality (q<20) were filtered using Trimmomatic program^16^, followed by removal of chimeric sequences via UCHIME program^17^. QIIME2 microbiome bioinformatics platform was utilized for downstream analysis of filtered reads^18^. Sequences with length shorter than 200 bases and ambiguous bases were removed by DADA2 denoising tool^19^. Greengenes and SILVA ribosomal sequence databases were utilized for taxonomic assignment through RDP classifier in QIIME2^20-22^.

Alpha diversity indices were calculated using Shannon-Weaver and Simpson’s reciprocal methods^23^, while beta diversity among the samples was measured according to weighted UniFrac distance matrices^24^. Extended error bar plots for differentially abundant taxa at phylum and genus level were constructed according to Welch’s t-test at 95% confidence interval with Storey’s FDR correction using STAMP statistical tool^25^.

## 3. Results

Oral examination of participants was carried out at the time of saliva collection. Figure 1 represents oral lesions in gukta chewers. Red erythroplakia of buccal mucosae, stains of gutka preparation and gutka deposition in dental plaque can be seen in the images.

**Figure 1.**
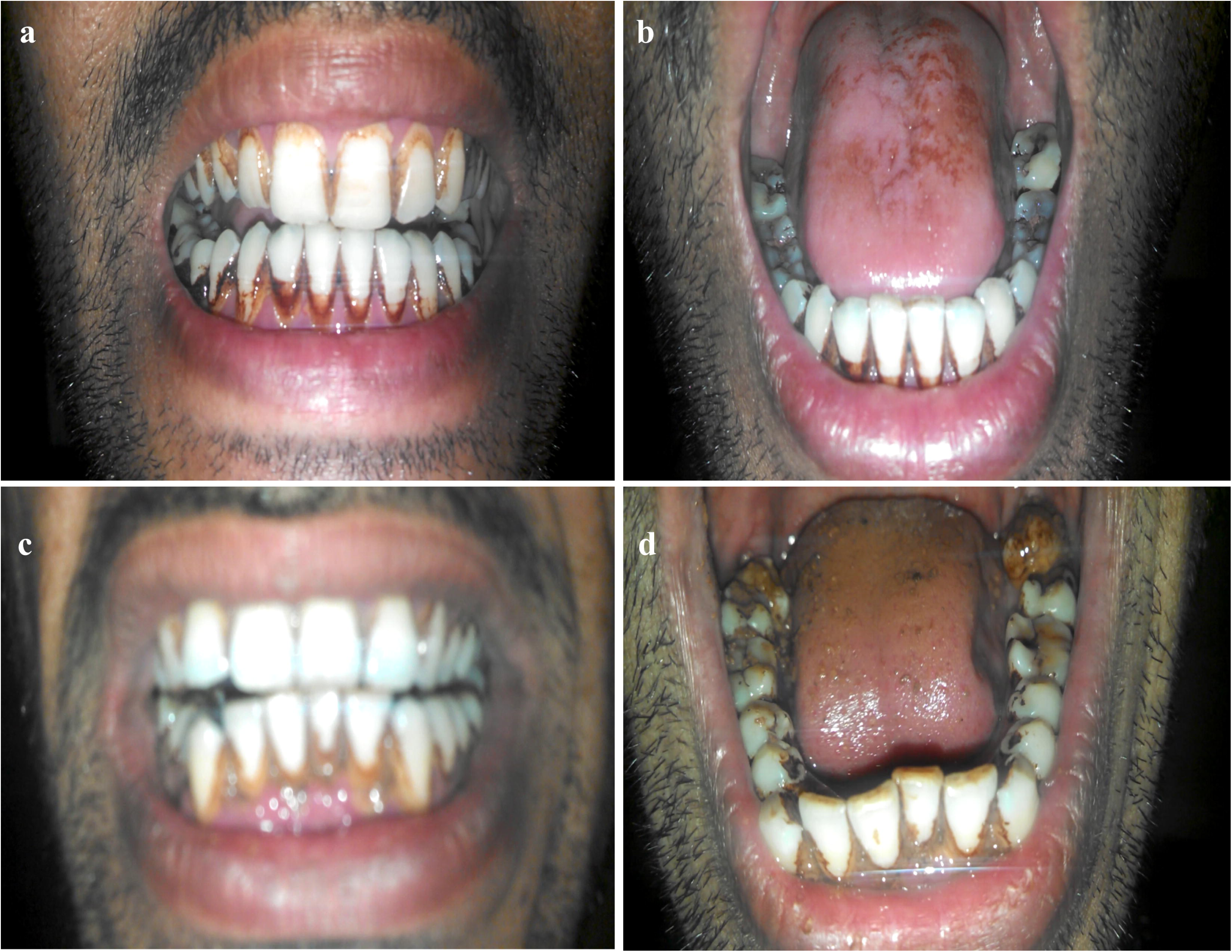
Oral cavity lesions in chewers of betel nut preparation “Gutka”.

### 3.1. 16S rDNA Sequencing and Assessment of Alpha Diversity

Filtration of low quality next-generation sequence (NGS) reads (i.e. q<20) resulted in 4,175,739 and 715,460 paired-end reads from salivary samples of non-chewers (n=55) and Gutka chewers (n=16), respectively. The read lengths of filtered NGS sequences were 250-300 nucleotides. Table 1 demonstrates the number of NGS reads and alpha diversity indices (Shannon-Weaver and Simpson’s methods) for both groups. The total number of genera in both groups was identified in order to evaluate bacterial diversity. Figure 2 demonstrates the VENN diagram of bacterial genera identified in Gutka chewers and non-chewers. In total, 234 and 75 bacterial genera were detected in non-chewers and Gutka chewers, respectively. Sixty-four genera were common between both groups, while 170 genera were unique for non-chewers and 11 genera were exclusively observed in Gutka chewers. Bacterial genera exclusively detected in gutka chewers were *Anaerophaga, Ancylomarina, Anoxybacillus, Bifidobacterium, Cellulosilyticum, Hyphomonas, Marinifilum, Mesocricetibacter, Pelomonas, Peptoniphilus, Scardovia*. Shannon-Weaver and Simpson’s alpha diversity indices represent variations in bacterial diversity within the samples^23^. On average, alpha diversity (Simpson’s reciprocal index) for saliva of non-chewers was found to be 11.0±4.6, while for Gutka chewers it was observed to be 7.27±3.03. Hence, the Gutka chewers demonstrated lower alpha diversity in comparison to non-chewers. NGS reads were assigned to their respective taxonomic groups in order to unravel the bacterial diversity trends amidst non-chewers and Gutka chewers. Welch’s t-test with Storey’s FDR correction at 95% confidence interval was applied to characterize significant variation of bacterial phyla amongst both groups (Figure 3a). No significant change in abundance of Bacteroidetes (p-value = 0.378) and Proteobacteria (p-value = 0.949) was observed in both groups, while sequences related to Firmicutes (p-value = 0.042) were significantly higher in Gutka chewers.

**Table 1:** Number of NGS reads, Simpson’s and Shannon-Weaver alpha diversity indices of Non-chewers (n=55) and Gutka chewers (n=16).

|  |  | Number of NGS reads | Simpson's reciprocal index | Shannon-Weaver index |
| --- | --- | --- | --- | --- |
| **Non-chewers** | Total | 4,175,739 | 604.76 | 229.09 |
|  | Average | 75,922 SD±43,448 | 11.0 SD±4.6 | 4.17 SD±0.6 |
| **Gutka chewers** | Total | 715,460 | 116.332 | 54.185 |
|  | Average | 44,716 SD±15,629 | 7.27 SD±3.03 | 3.38 SD±0.7 |

**Figure 2.**
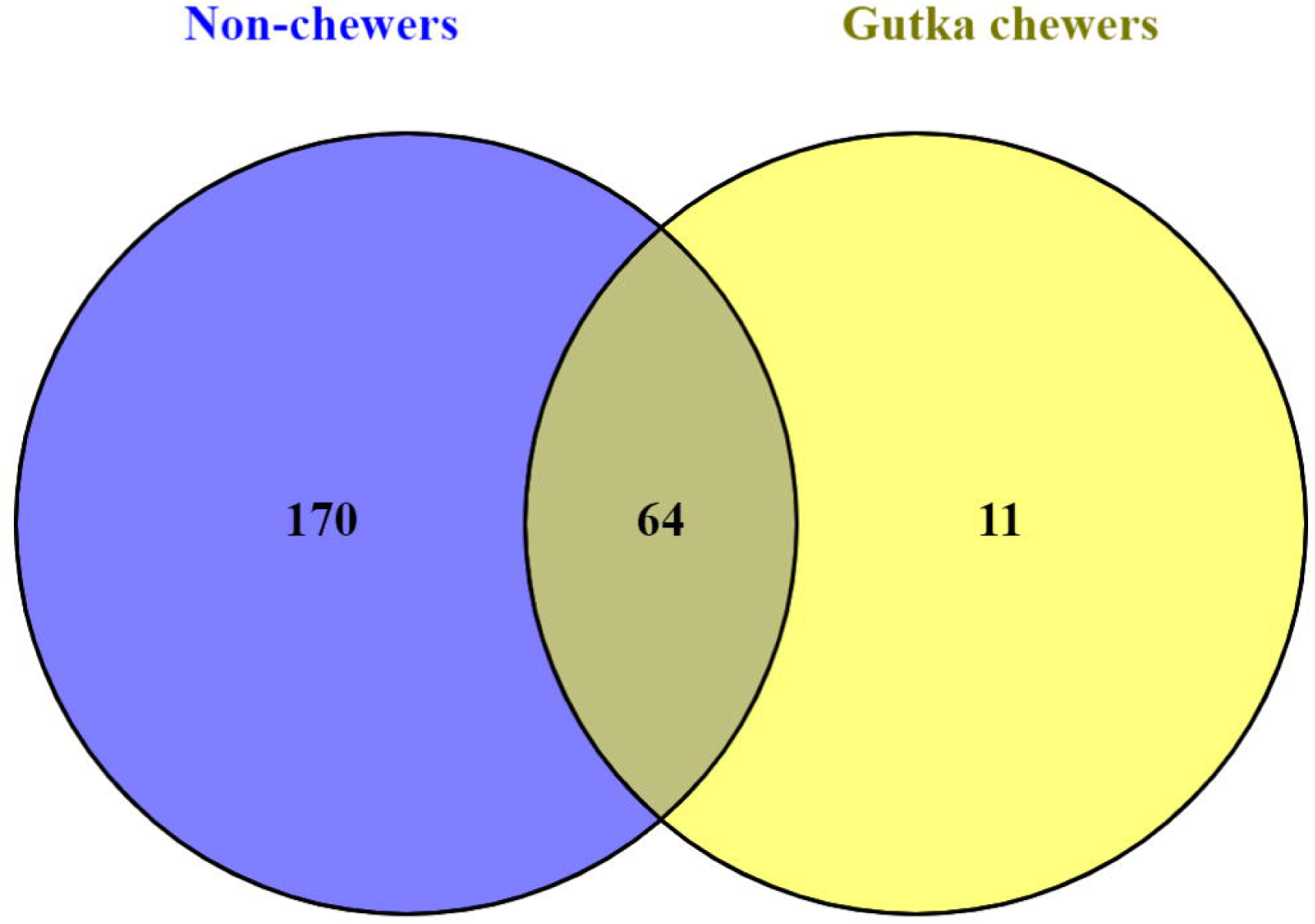
VENN diagrammatic plot representing unique and shared salivary bacterial genera in non-chewers and Gutka chewers.

**Figure 3.**
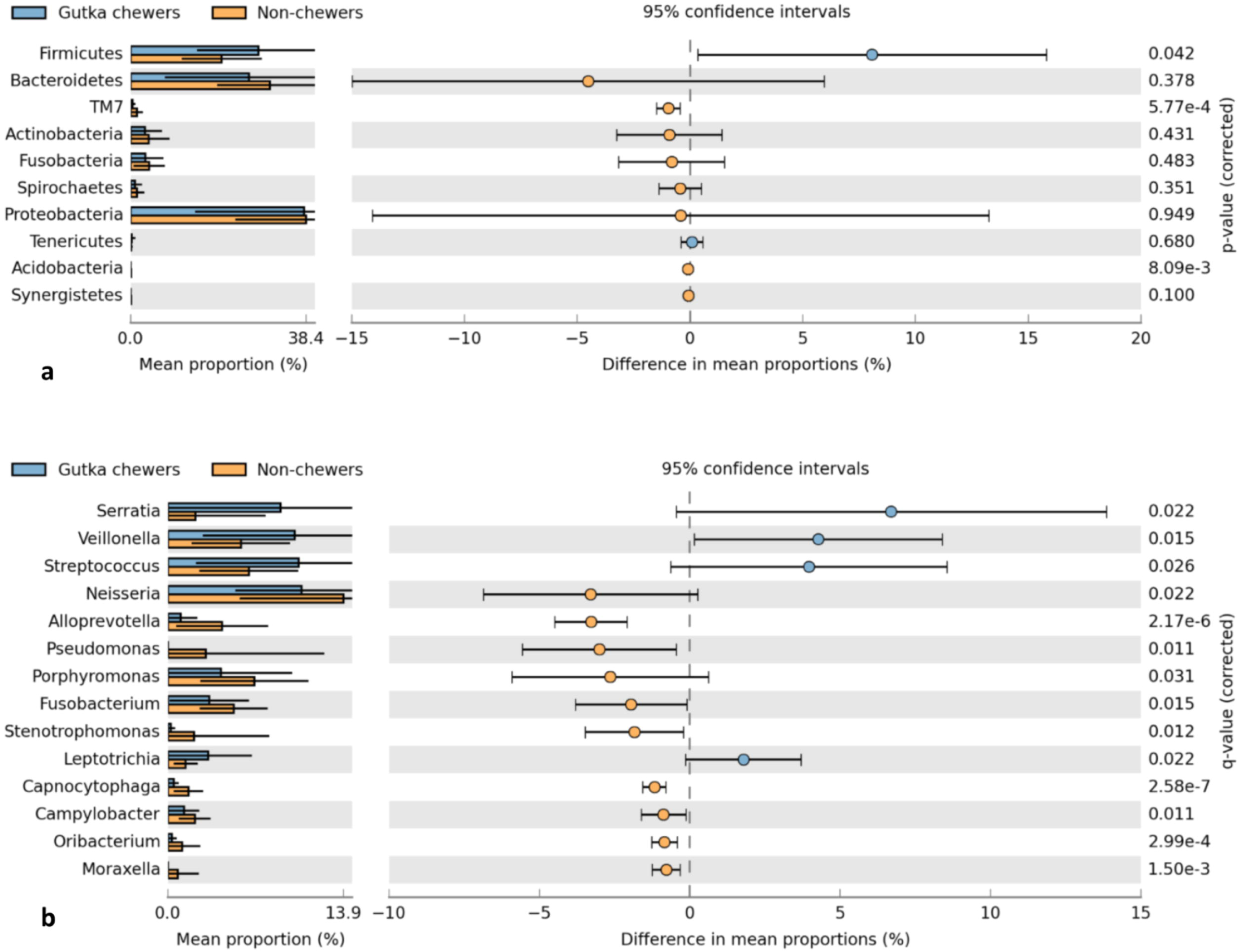
(a) Extended error bar plot representing Welch’s t-test based differential abundance profile of bacterial phyla in non-chewers (n = 55) and Gutka chewers (n = 16) at 95% confidence interval with Storey’s FDR correction. (b) Extended error bar plot representing Welch’s t-test based differential abundance profile of bacterial genera in non-chewers (n = 55) and Gutka chewers (n = 16) at 95% confidence interval with Storey’s FDR correction.

### 3.2. Bacterial Diversity at Genus Level

Bacterial diversity at genus level was measured by weighted UniFrac distance matrix (WUDM) to determine the beta diversity patterns amongst Gutka chewers and non-chewers. Weighted UniFrac distance matrix calculates the diversity among the samples based on the differentially abundant taxa^24^. The data generated by WUDM was utilized to construct PCoA plot. In the PCoA plot, samples from non-chewers clustered together, while samples from Gutka chewers scattered in two regions (figure 4). Samples of Gutka chewers remained divergent from the non-chewers cluster, which is an indicator of dissimilarity of bacterial diversity between both groups.

**Figure 4.**
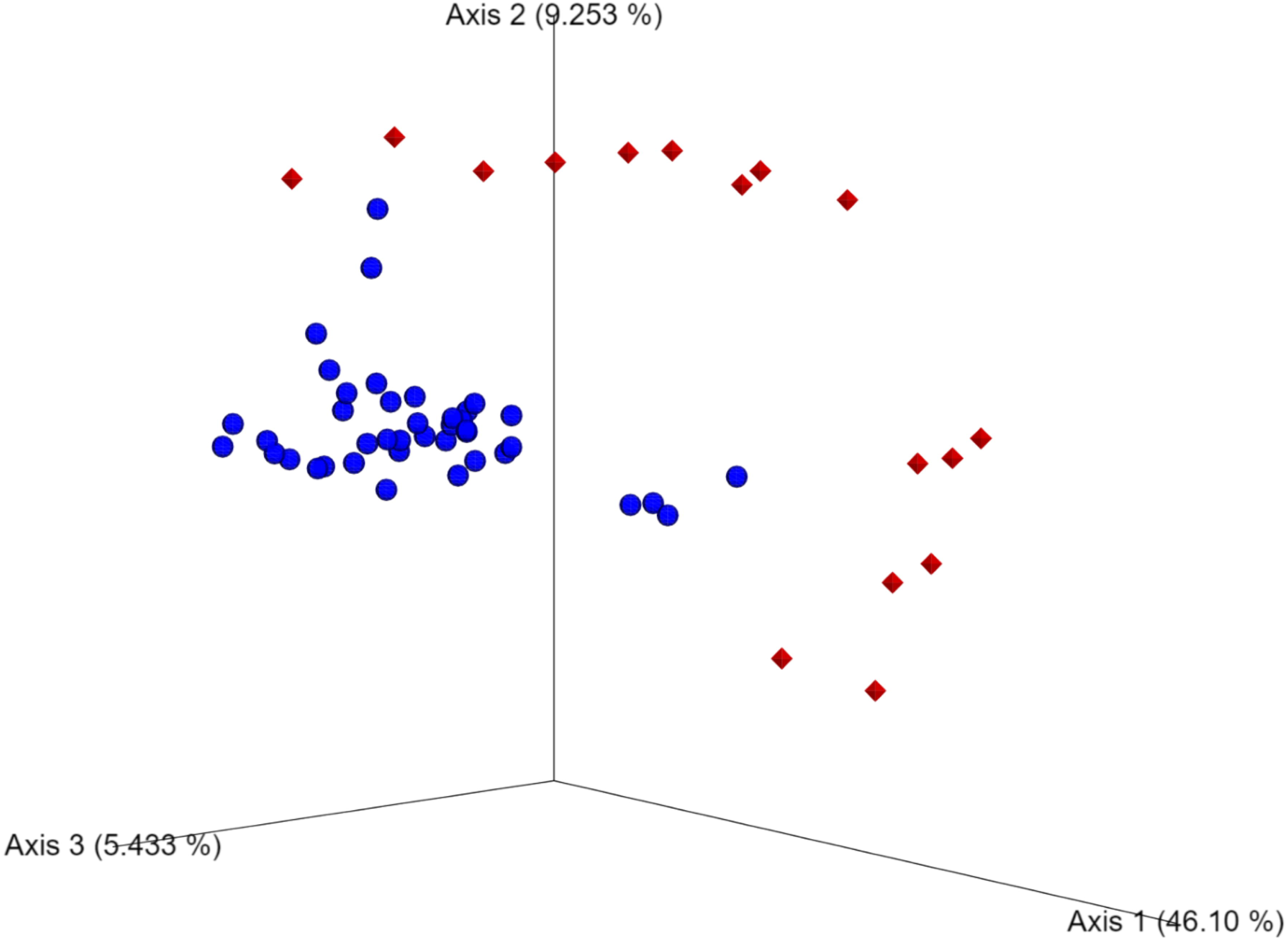
Weighted uniFrac distance based PCoA plot of samples from non-chewers (n = 55) and Gutka chewers (n = 16). ♦ and ⋅ represent samples of Gutka chewers and non-chewers, respectively.

To characterize the statistical abundance profile of bacterial genera in both groups Welch’s t-test with Storey’s FDR correction (95% confidence interval) was applied. The abundance of 4 bacterial genera was found to be significantly elevated in Gutka chewers (Figure 3b). These bacterial genera were *Serratia* (p-value = 0.022), *Veillonella* (p-value = 0.015), *Streptococcus* (p-value = 0.026) and *Leptotrichia* (p-value = 0.022). Whereas, the population of four bacterial genera decreased in Gutka chewers which were *Neisseria* (p-value = 0.022), *Alloprevotella* (p-value = 2.16 X 10^−6^), *Pseudomonas* (p-value = 0.011) and *Fusobacterium* (p-value = 0.015).

## 4. Discussion

Oral cavity provides a microenvironment for the growth of hundreds of bacterial species^11^. These oral bacterial communities play a role in sustainability of normal oral homeostatic state. However, many intrinsic and environmental factors could stimulate microbial changes and lead to dysbiosis^26^.

In the present study, the salivary microbiome of chewers of betel nut preparations “Gutka” (n=16) and non-chewers (n=55) was analyzed by using 16S rDNA metagenomics approach. Both alpha diversity indices (i.e. Shannon-Weaver and Simpson’s) appeared to be lower for the betel nut addict individuals in comparison to non-addicts (table 1). Furthermore, before number of unique bacterial genera were also found to be lower for gutka chewers (figure 2), which is suggestive of decrease in oral microbiome diversity in response to gutka chewing. Among the body sites, oral cavity is known to possess diverse bacterial population as indicated by higher alpha diversity values^27^. In the present study, we found decreased diversity of bacterial communities in saliva of gutka chewers in comparison to non-chewers. Recently, it has been reported that decrease in alpha diversity is correlated with onset and progression of dental caries^28^.

Among the bacterial phyla, Firmicutes were found to be significantly higher in abundance in gutka chewers than in non-chewers (Figure 3a). These findings are in support of a previous study, which reported similar oral microbial patterns in patients suffering from metabolic syndrome^29^. Furthermore, abundance of oral Firmicutes population is also previously reported in correlation with elevated levels of inflammation^30^. In a previous study from USA, Actinobacteria phylum was depicted to be second most abundant oral bacterial phylum associated with betel nut chewing^15^. In contrast, our results did not indicate any significant correlation of Actinobacteria population in response to consumption of betel nut preparation (Gutka).

Beta diversity measurement (Weighted uniFrac Distance Matrix) demonstrated that both groups are in separate clusters indicating substantial variations in salivary microbiome on the genus level (Figure 4). The abundance of Serratia (p-value = 0.022), Veillonella (p-value = 0.015), Streptococcus (p-value = 0.026) and Leptotrichia (p-value = 0.022) bacterial genera was found to be significantly associated with gutka chewers (Figure 3b). Our results are in agreement with a previous study which by using denaturing gradient gel electrophoresis (DGGE) correlated the abundance of Streptococcus and Veillonella in response to usage of betel nut preparations^31^. Serratia species are gram-negative, facultatively anaerobic bacteria possessing the ability to act as opportunistic pathogens in immunocompromised patients. These bacteria are found in oral cavities of patients suffering from chronic periodontitis^32^. Veillonella species are strictly anaerobic, biofilm-producing and aciduric bacteria that thrive in correlation with acidogenic bacterial species such as Streptococci^33^. Acidogenic species such as Streptococci and Leptotrichia dissimilate salivary disaccharides into simpler monosaccharides (i.e. glucose) followed by conversion of glucose into organic acids (i.e. lactate), which in turn is then utilized by Veillonella species as the energy and carbon source for growth^34,33^. Furthermore, Veillonella species can adhere to teeth and gums by using dextran produced by Streptococci through the action of their glucosyltransferases on sucrose^35^. Leptotrichia species are facultatively anaerobic, acidogenic bacteria which act as causative agents of oral lesions and dental caries in immunocompromised patients^34^. A consortium of these acidogenic and aciduric bacterial genera in Gutka chewers may contribute in deterioration of oral and dental health.

The present study provides additional information related to oral microbial dysbiosis in Gutka chewers, which might be helpful in further assessment of oral complications that arise due to consumption of betel nut preparations.

## Data Availability

All data related to this study would be available on request.

## Acknowledgments

We thank all participants of this study.

## Conflict of interest

Authors declare no conflict of interest.

## Notes

### Competing Interest Statement

The authors have declared no competing interest.

### Clinical Trial

The study was sanctioned by Independent Ethics Committee, International Center for Chemical and Biological Sciences, University of Karachi, Pakistan (Study no. 011-SS-2016. Protocol no. ICCBS/IEC-011-SS-2016/Protocol/1.0).

### Funding Statement

Funding for this study was provided by the International Center for Chemical and Biological Sciences, University of Karachi, Pakistan.

